# Low serum vitamin D level and COVID-19 infection and outcomes, a multivariate meta-analysis

**DOI:** 10.1101/2020.10.24.20218974

**Authors:** Jie Chen, Lixia Xie, Ping Yuan, Jianyong Ma, Peng Yu, Chunhua Zheng, Xiao Liu

## Abstract

**Objective:** This study aimed to determine whether serum vitamin D is independently associated with COVID-19 infection and outcomes in patients with COVID-19.

**Methods:** We identified relevant studies by searching the PubMed, Embase, and medRxiv databases from December 2019 to October 1, 2020. Odds ratios (ORs) were pooled using random-effects models. Only reports with multivariate adjusted results were included to avoid the impact of potential confounding factors.

**Results:** A total of six studies with 377,265 patients were identified. Overall, in the categorical analysis, a low serum vitamin D level was associated with an increased risk of COVID-19 infection (OR: 1.47, 95% CI: 1.09- 1.97, I2=81%), hospitalization (OR: 1.83, 95% CI: 1.22-2.74, I2=0%), but not in-hospital death (OR: 2.73, 95% CI: 0.27-27.61). Notably, when vitamin D level was analyzed as a continuous variable, each 5 ng/ml increase in vitamin D level was not associated with any increased risk of COVID-19 infection (OR: 1.04, 95% CI: 0.96-1.12, I2=74%) or in-hospital death (OR: 1.02, 95% CI: 0.93-1.12).

**Conclusions:** Low serum vitamin D is associated with an increased risk of COVID-19 infection and hospitalization. In-hospital death showed a tendency to be increased in COVID-19 patients with low vitamin D levels. The ongoing clinical trials for evaluation of vitamin D supplementation will be key to the validation of this adjunctive treatment for COVID-19 patients.

## Introduction

Recent studies have highlighted that mean plasma vitamin D level is significantly lower among those who tested positive for COVID-19 than among those who tested negative[1]. Rhodes et al.[2] found that all countries that lie below 35 degrees north latitude have relatively low mortality for patients with COVID-19. These results suggest that low levels of vitamin D may be associated with increased COVID-19 infection rates and worse outcomes in COVID-19. Consistently, several studies have reported that serum vitamin D deficiency is associated with an increased risk of COVID-19 positivity and worse outcomes (e.g., severe COVID-19 and in-hospital death)[3-5]. For example, in Radujkovic’s cohort (N=198), vitamin D deficiency was associated with a higher risk of invasive mechanical ventilation and/or in-hospital death (HR: 6.12, 95% CI: 2.79–13.42 and HR: 14.73, 95% CI: 4.16–52.19, respectively)[5]. However, a larger cohort-based study from the UK Biobank showed a positive association between serum vitamin D concentration and severe COVID-19 infection (per 10 nmol/L vitamin D, HR: 0.93; p<0.001) and mortality (per 10 vitamin D nmol/L, HR: 0.92; p=0.016) in univariate analysis, but not after adjustment for confounders (COVID-19 infection HR: 1.00; p=0.89; mortality HR: 0.98; p=0.69)[6]. Similarly, two other studies also found no association between vitamin D level and COVID-19 positivity[7, 8]. Therefore, the impact of vitamin D on COVID-19 is still controversial, and a definite conclusion has not yet been drawn. We therefore aimed to conduct a meta-analysis to clarify the association between serum low vitamin D and COVID-19.

## Methods

This study was performed according to Preferred Reporting Items for Systematic reviews and Meta-Analyses Statement (PRISMA) guidelines.

### Literature Search and study Selection

Two authors independently searched several databases (PubMed, Embase, medRxiv) using the following two groups of keywords with no language restrictions: 2019-novel coronavirus, SARS-CoV-2, COVID-19, 2019-nCoV, and vitamin D. In addition, we searched the reference lists of other relevant publications to identify further studies. Inclusion criteria were as follows: 1) human studies, not laboratory or animal studies, that were published as original articles; 2) case-control study or cohort study; 3) reports with multivariate analyses with estimate effect (odds ratio) and its 95% confidence intervals (CIs) results that reported vitamin D and COVID-19 (e.g., COVID-19 infection risk, severity, and in-hospital death). When multiple papers reporting on the same study were identified, the most informative or complete article was included.

### Data extraction and statistical analysis

Two authors independently extracted all data from included studies. In cases of discrepancies, disagreement was resolved by collegial discussion. The following information was extracted: first author, country, publication year, gender, mean or median age, study design, sample size, vitamin D level, OR or RR with the 95% CI for each category (results adjusted according to most potential confounders), and adjusted variables.

Odds ratios (ORs) were pooled using random-effects models. We calculated study-specific slopes (vitamin D per 5 ng/ml decrement) and 95% CIs from the natural logs of the reported RRs and CIs across categories of vitamin D level[9, 10]. Cochran Q and I2 statistics were used to detect statistical heterogeneity between studies. The overall quality of included studies was assessed by Newcastle-Ottawa quality assessment scale (NOS), with a NOS score of ≥6 stars was regarded as high-quality[11]. All statistical analyses were conducted by using Review Manager version 5.3 (The Cochrane Collaboration 2014; Nordic Cochrane Center Copenhagen, Denmark). All statistical tests were double-sided, and P < 0.05 was considered statistically significant.

## Results

### Study selection

As shown in **Figure 1**, we identified 486 studies in the initial database search. After removing duplicates(n = 257) and studies with insufficient information on vitamin D and COVID-19, a total of six[3-6, 12, 13] studies with 377,265 patients remained.

**Figure 1.**
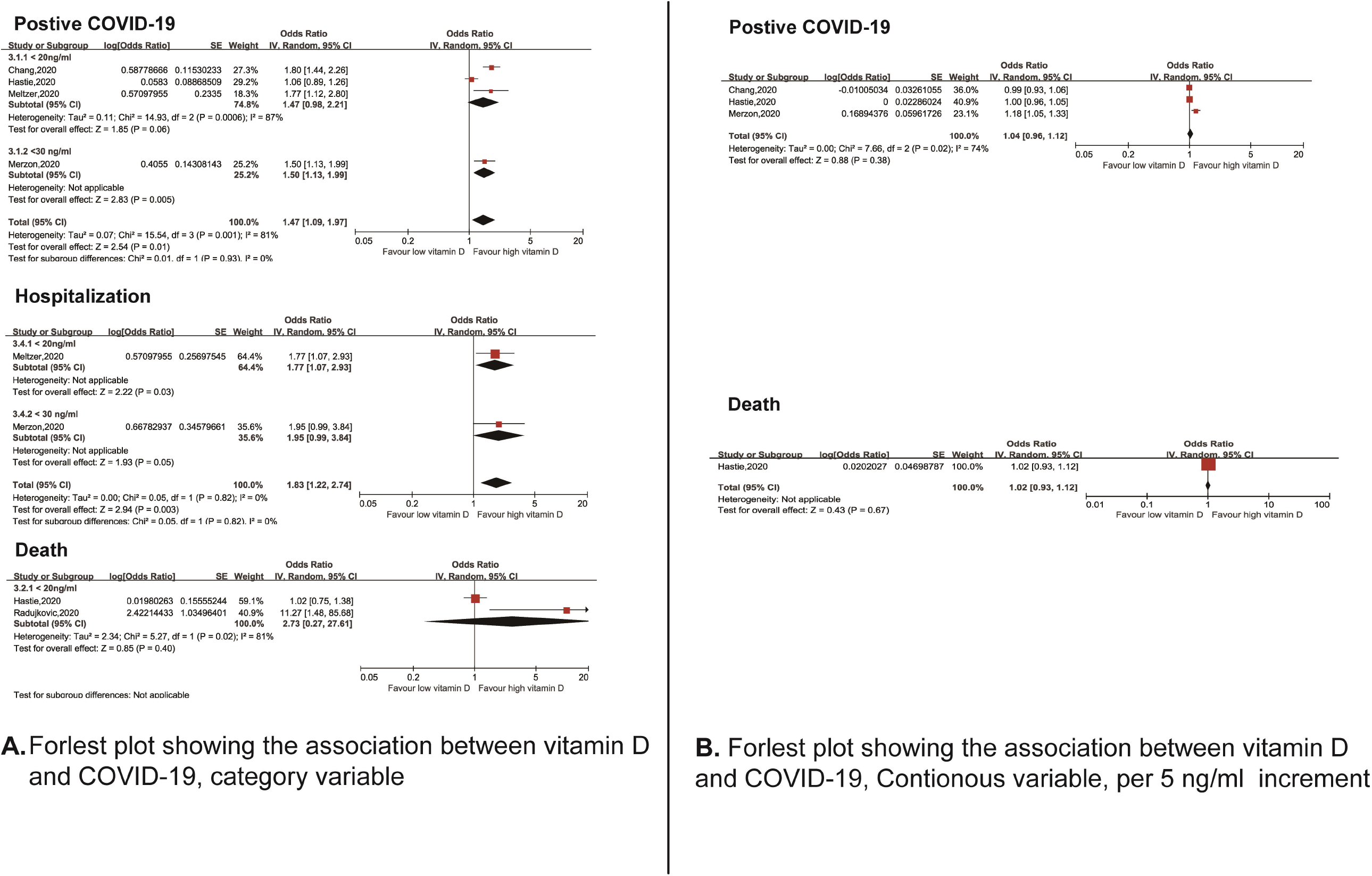
Forest plot showing the association between serum vitamin D level and risk of COVID-19 positivity, hospitalization, and in-hospital death in patients with COVID-19. **A:** Vitamin D analyzed as a categorical variable; **B:** Vitamin D analyzed as a continuous variable (per 5 ng/ml decrement).

### Study characteristics and quality

Each study is listed in **Table 1**. Four[3, 6, 12, 13] studies reported vitamin D levels and COVID-19 positivity, and four[3, 5, 6, 12] reported the association of vitamin D levels and COVID-19 outcomes. Of the six studies, 3 were cohort studies and 3 were case-control cohort studies. Most of studies (n = 3) were performed in USA, 1 was performed in UK, 1 in Israel and 1 in Germany. The overall quality of included studies was high (NOS score ≥ 6)(**Table 1**).

**Table 1.**
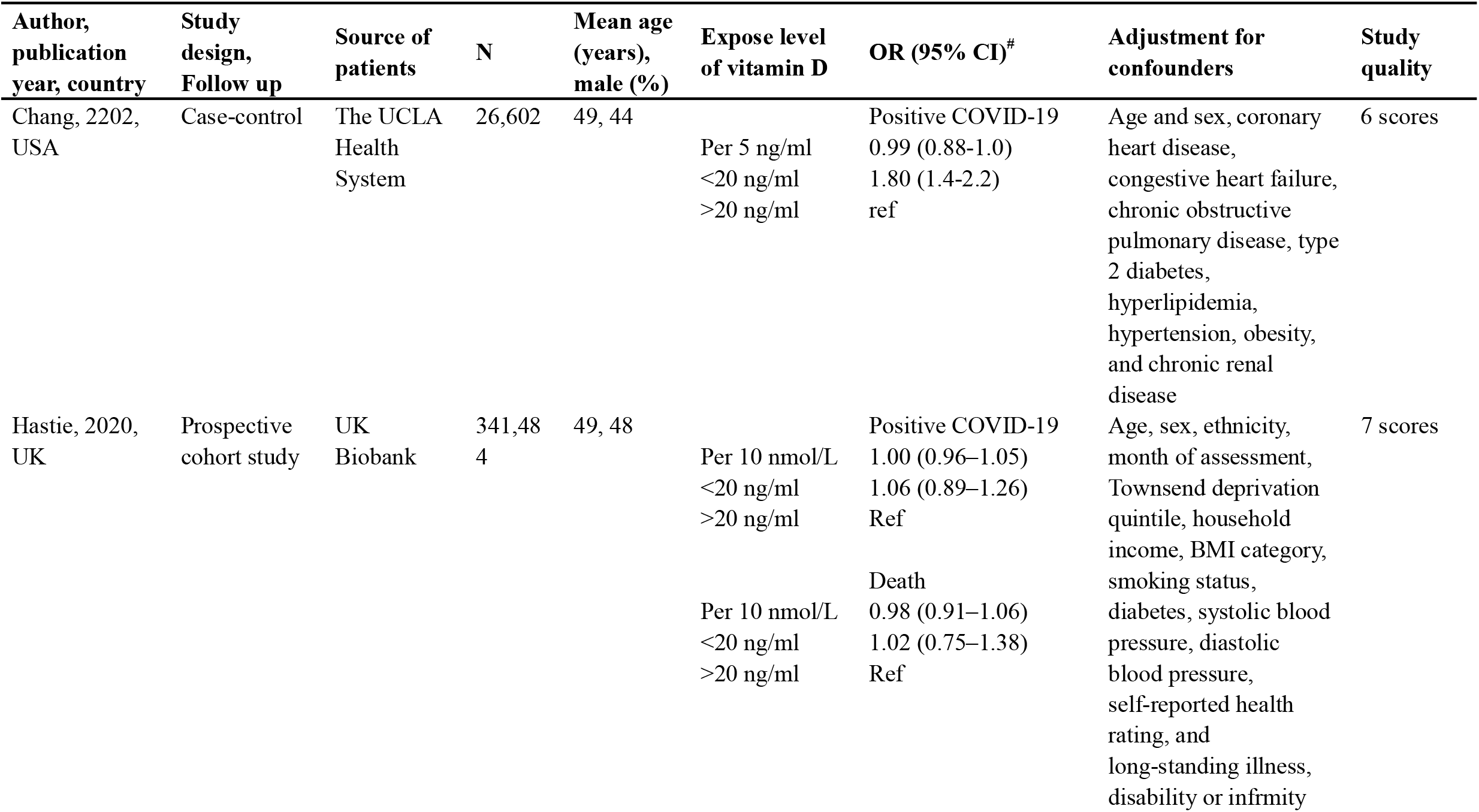
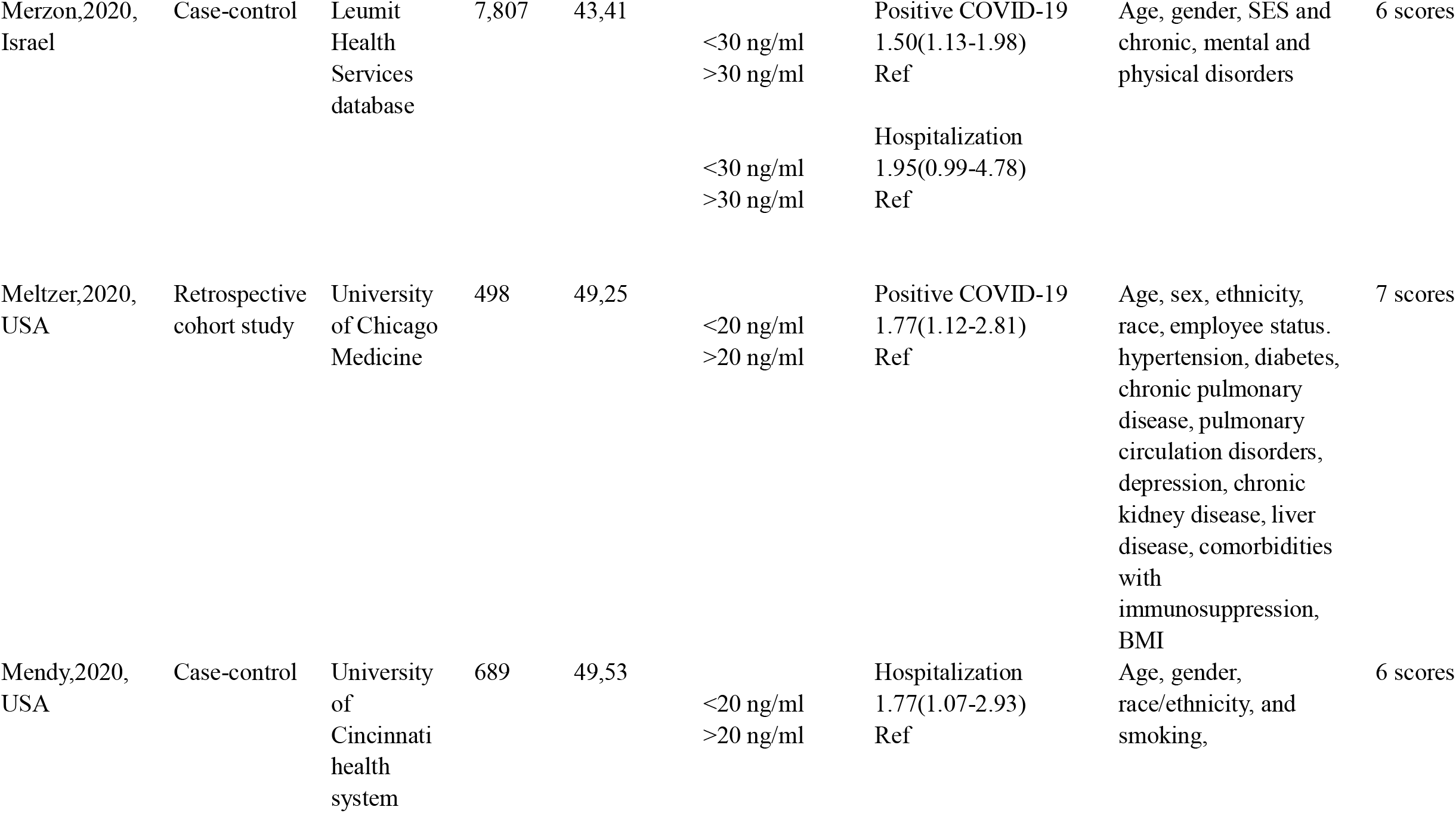
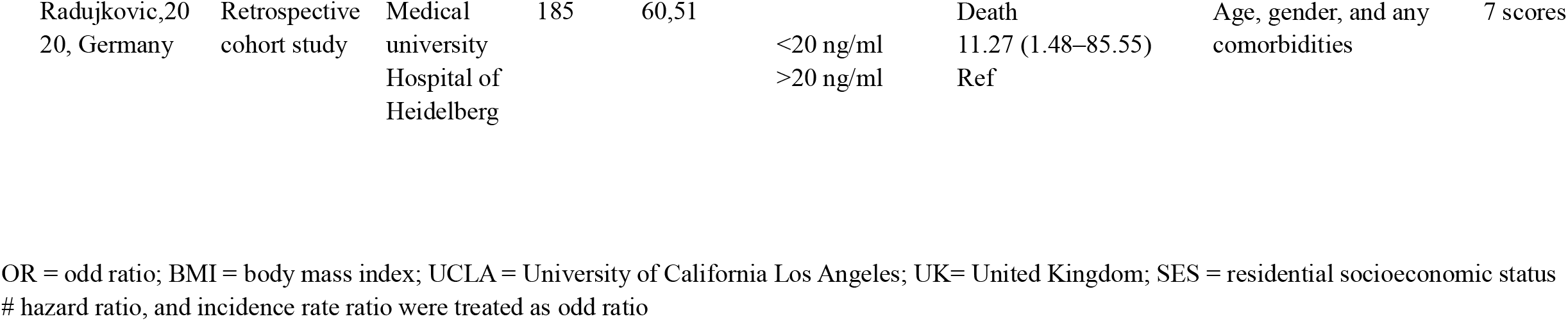
Basic characteristics of included articles in the meta-analysis.

| Author, publication year, country | Study design, Follow up | Source of patients | N | Mean age (years), male (%) | Expose level of vitamin D | OR (95% CI)<sup>#</sup> | Adjustment for confounders | Study quality |
| --- | --- | --- | --- | --- | --- | --- | --- | --- |
| Chang, 2202, USA | Case-control | The UCLA Health System | 26,602 | 49, 44 | Per 5 ng/ml<br><20 ng/ml<br>>20 ng/ml | Positive COVID-19<br>0.99 (0.88-1.0)<br>1.80 (1.4-2.2)<br>ref | Age and sex, coronary heart disease, congestive heart failure, chronic obstructive pulmonary disease, type 2 diabetes, hyperlipidemia, hypertension, obesity, and chronic renal disease | 6 scores |
| Hastie, 2020, UK | Prospective cohort study | UK Biobank | 341,484 | 49, 48 | Per 10 nmol/L<br><20 ng/ml<br>>20 ng/ml<br>Per 10 nmol/L<br><20 ng/ml<br>>20 ng/ml | Positive COVID-19<br>1.00 (0.96–1.05)<br>1.06 (0.89–1.26)<br>Ref<br>Death<br>0.98 (0.91–1.06)<br>1.02 (0.75–1.38)<br>Ref | Age, sex, ethnicity, month of assessment, Townsend deprivation quintile, household income, BMI category, smoking status, diabetes, systolic blood pressure, diastolic blood pressure, self-reported health rating, and long-standing illness, disability or infirmity | 7 scores |
| Merzon,2020,<br>Israel | Case-control | Leumit<br>Health<br>Services<br>database | 7,807 | 43,41 | <30 ng/ml<br>>30 ng/ml | Positive COVID-19<br>1.50(1.13-1.98)<br>Ref | Age, gender, SES and<br>chronic, mental and<br>physical disorders | 6 scores |
|  |  |  |  |  | <30 ng/ml<br>>30 ng/ml | Hospitalization<br>1.95(0.99-4.78)<br>Ref |  |  |
| Meltzer,2020,<br>USA | Retrospective<br>cohort study | University<br>of Chicago<br>Medicine | 498 | 49,25 | <20 ng/ml<br>>20 ng/ml | Positive COVID-19<br>1.77(1.12-2.81)<br>Ref | Age, sex, ethnicity,<br>race, employee status.<br>hypertension, diabetes,<br>chronic pulmonary<br>disease, pulmonary<br>circulation disorders,<br>depression, chronic<br>kidney disease, liver<br>disease, comorbidities<br>with<br>immunosuppression,<br>BMI | 7 scores |
| Mendy,2020,<br>USA | Case-control | University<br>of<br>Cincinnati<br>health<br>system | 689 | 49,53 | <20 ng/ml<br>>20 ng/ml | Hospitalization<br>1.77(1.07-2.93)<br>Ref | Age, gender,<br>race/ethnicity, and<br>smoking, | 6 scores |
| Radujkovic,20<br>20, Germany | Retrospective<br>cohort study | Medical<br>university<br>Hospital of<br>Heidelberg | 185 | 60,51 | <20 ng/ml<br>>20 ng/ml | Death<br>11.27 (1.48–85.55)<br>Ref | Age, gender, and any<br>comorbidities | 7 scores |

OR = odd ratio; BMI = body mass index; UCLA = University of California Los Angeles; UK= United Kingdom; SES = residential socioeconomic status # hazard ratio, and incidence rate ratio were treated as odd ratio
OR = odd ratio; BMI = body mass index; UCLA = University of California Los Angeles; UK= United Kingdom; SES = residential socioeconomic status

### The effect of vitamin D level on COVID-19

Overall, in the categorical analysis, low serum vitamin D level was associated with increased risk of COVID-19 infection (OR: 1.47, 95% CI: 1.09-1.97, I2=81%), hospitalization (OR: 1.83, 95% CI: 1.22-2.74, I2=0%) but not in-hospital death (OR: 2.73, 95% CI: 0.27-27.61) (**Figure 1.A)**. When vitamin D level was analyzed as a continuous variable, each 5 ng/ml increase in vitamin D level was not associated with an increased risk of COVID-19 positivity (OR: 1.04, 95% CI: 0.96-1.12, I2=74%) or in-hospital death (OR: 1.02, 95% CI: 0.93-1.12) (**Figure 1.B**). The publication bias of POAF was not conducted as limited studies (N<10) according to the guidelines [14].

## Discussion

To the best of our knowledge, this is the first meta-analysis to investigate the association between serum vitamin D and COVID-19 positivity rates and outcomes in patients with COVID-19. Our results showed that low vitamin D independently increased the risk of COVID-19 infection and hospitalization by 47% and 83%, respectively. However, we did not find a significant positive association between low vitamin D and in-hospital death (P=0.56). Considering the limited number of studies in the present study, more studies with a larger sample size are needed to verify these results. Several studies have found a positive correlation between mean serum vitamin D levels and the number of in-hospital deaths caused by COVID-19 cases and mortality in European countries[2, 15, 16]. In another study with a small sample size (N=46), a survival analysis outlined that patients with severe vitamin D deficiency (<10 ng/ml) had a 50% increase in mortality probability compared with patients with vitamin D >10 ng/ml[17]. Furthermore, Mendy et al.[4] also found that vitamin D increased the risk of severe COVID-19 risk and ICU/in-hospital death in multivariate analysis. Another study reported that vitamin D insufficiency increased the risk of IMV/in-hospital death by 5.75-fold after adjusting for age, gender, and comorbidities[5]. Therefore, whether vitamin D insufficiency increases the severity of the disease and consequently the risk of in-hospital death is still controversial. More studies adjusted for clinical cofounders are needed to clarify this issue in future research.

Notably, we found that the results for vitamin D level analyzed as a categorical variable and as a continuous variable are inconsistent. Apart from the limited number of studies included in the continuous analysis, we speculate that this might be explained by a nonlinear dose-response relationship between serum vitamin D and COVID-19. Such a nonlinear dose-response relationship between vitamin D and disease susceptibility/severity is common[18-20], reflecting a significant difference in the effects of high vitamin D and low vitamin D levels on COVID-19. Therefore, a linear model (continuous variable) may be unable to actually detect the impact of low vitamin D on COVID-19.

There are several potential underlying pathophysiological mechanisms for the association between serum vitamin D deficiency and COVID-19. First, various studies have shown the antiviral activities of vitamin D through effects on dendritic cells and T cells[21]. A meta-analysis of randomized controlled trials also demonstrated that vitamin D treatment for patients with vitamin D deficiency can reduce viral respiratory infections, among which coronaviruses are common causative organisms[22]. Another vital mechanism might be the inhibition of the renin-angiotensin-aldosterone system (RAAS). Our previous meta-analyses have suggested that ACEI/ARB treatment by inhibiting RAAS might reduce COVID-19 infection and improve outcomes in patients with COVID-19[23]. Vitamin D could inhibit AT-1R and stimulate ACE2, thus inhibiting inflammatory responses, including limiting the inflammatory cytokine storm (e.g., IL-6, TNFα)[24].

Based on the positive association between vitamin D deficiency and COVID-19, it is reasonable to suggest that vitamin D supplementation might reduce the risk of COVID-19 infection and improve outcomes in patients with COVID-19. The benefits of vitamin D supplementation in people who are vitamin D deficient are known to include reducing other viral respiratory infections (e.g., respiratory syncytial virus and influenza infection). Furthermore, a meta-analysis of individual participant data also showed that vitamin D supplementation substantially reduced the severity of COPD[25]. Therefore, we hypothesize that vitamin D supplementation might be useful to reduce the incidence and burden of COVID-19. However, there are no published articles that assess the effect of vitamin D supplements on COVID-19. Ongoing clinical trials (e.g., NCT04407286 and NCT04482673) for the evaluation of vitamin D supplementation will be key to the validation of this adjunctive treatment for COVID-19 patients[24].

### Study Limitations

The present meta-analysis has several limitations. There is high heterogeneity in our meta-analysis, which might derive from the study design and variability in baseline characteristics. For example, several studies have shown that the mortality rate for black patients is higher than the rate for white COVID-19 patients[26, 27]. Second, many factors can modulate vitamin D status, including genetic polymorphisms, age, health, sun exposure behavior, season, and so on[21]. Although we only included studies that performed multivariable analysis, some potential risk factors were not fully adjusted, which might also have affected our results. Therefore, further research might adjust for additional confounding factors.

## Conclusion

Low serum vitamin D is associated with an increased risk of COVID-19 infection and hospitalization. In-hospital death showed a tendency to be increased in COVID-19 patients with low vitamin D levels; however, this trend did not reach statistical significance, which might be due to the small sample size. Further larger studies controlling for confounding factors are needed to investigate the association between low vitamin D and COVID-19 related death. The ongoing clinical trials for evaluation of vitamin D supplementation will be key to the validation of this adjunctive treatment for COVID-19 patients.

## Supporting information

PRISMA

## Data Availability

all data was provided in mansucipt

## Funding information

This work was supported in part by the National Natural Science Foundation of China (No.81760050, 81760048) and the Jiangxi Provincial Natural Science Foundation for Youth Scientific Research (No. 20192ACBL21037).

### Competing financial interests

The authors declare no competing financial interests.

### AUTHORSHIP

Guarantor of the article: X.L.

## Author contributions

X.L contributed to study concept and design. M-L.L and J.C contributed to study research, acquisition of data and statistical analysis. All the authors reviewed and approved the final manuscript.

## Abbreviations

COVID-19: coronavirus disease 2019
OR: odds ratio
CI: confidence interval
IV: inverse variance
SE: standard error

---

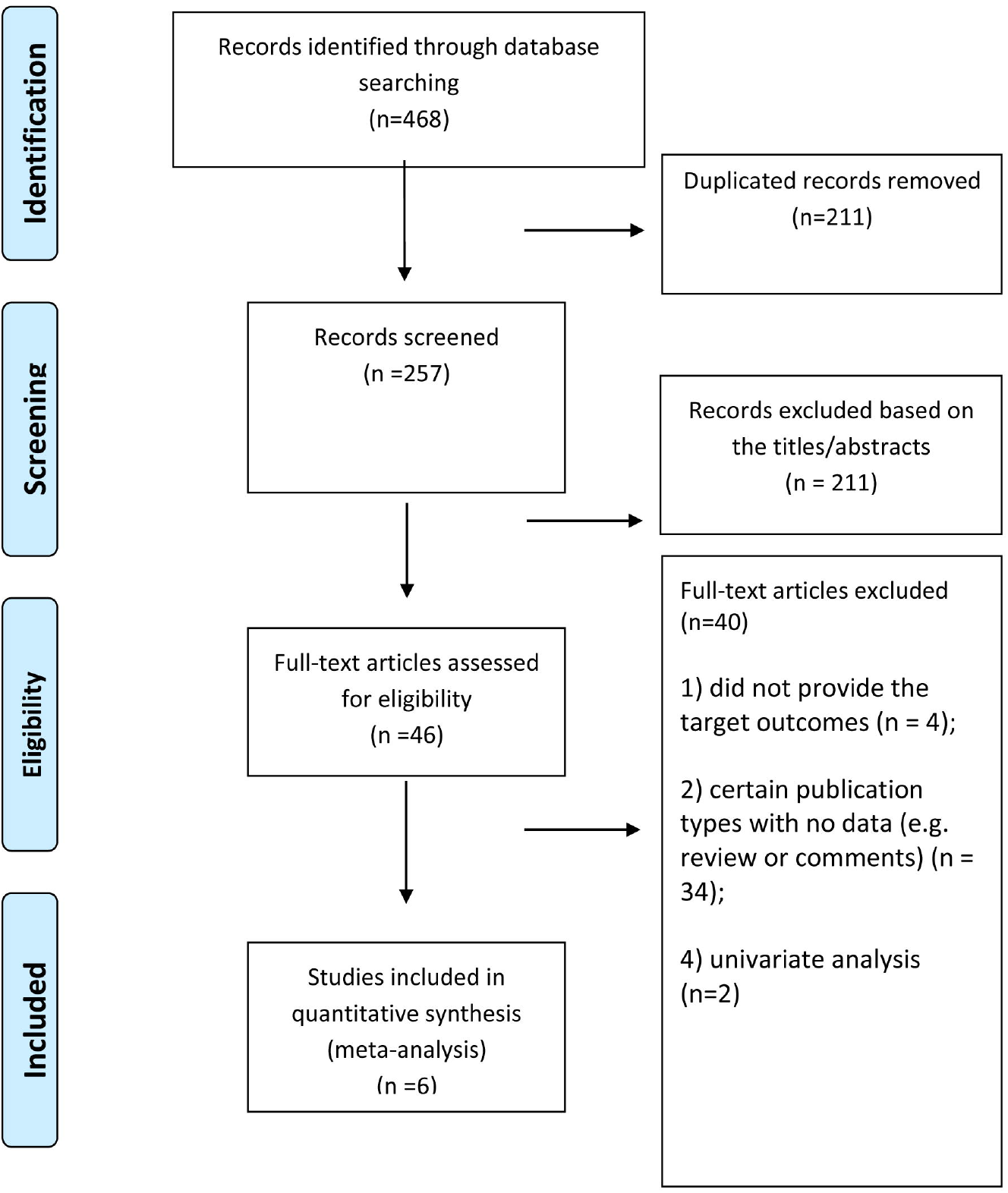

## Notes

### Competing Interest Statement

The authors have declared no competing interest.

## References

[1] Ali N. Role of vitamin D in preventing of COVID-19 infection, progression and severity. J Infect Public Health. 2020;13:1373–80.

[2] Rhodes JM, Subramanian S, Laird E, Kenny RA. Editorial: low population mortality from COVID-19 in countries south of latitude 35 degrees North supports vitamin D as a factor determining severity. Aliment Pharmacol Ther. 2020;51:1434–7.

[3] Meltzer DO, Best TJ, Zhang H, Vokes T, Arora V, Solway J. Association of Vitamin D Status and Other Clinical Characteristics With COVID-19 Test Results. JAMA Netw Open. 2020;3:e2019722.

[4] Mendy A, Apewokin S, Wells AA, Morrow AL. Factors Associated with Hospitalization and Disease Severity in a Racially and Ethnically Diverse Population of COVID-19 Patients. medRxiv. 2020.

[5] Radujkovic A, Hippchen T, Tiwari-Heckler S, Dreher S, Boxberger M, Merle U. Vitamin D Deficiency and Outcome of COVID-19 Patients. Nutrients. 2020;12.

[6] Hastie CE, Pell JP, Sattar N. Vitamin D and COVID-19 infection and mortality in UK Biobank. Eur J Nutr. 2020.

[7] Raisi-Estabragh Z, McCracken C, Bethell MS, Cooper J, Cooper C, Caulfield MJ, et al. Greater risk of severe COVID-19 in Black, Asian and Minority Ethnic populations is not explained by cardiometabolic, socioeconomic or behavioural factors, or by 25(OH)-vitamin D status: study of 1326 cases from the UK Biobank. J Public Health (Oxf). 2020;42:451–60.

[8] Hastie CE, Mackay DF, Ho F, Celis-Morales CA, Katikireddi SV, Niedzwiedz CL, et al. Vitamin D concentrations and COVID-19 infection in UK Biobank. Diabetes Metab Syndr. 2020;14:561–5.

[9] Xu C, Thabane L, Liu T-Z, Li L, Borhan S, Sun X. Flexible piecewise linear model for investigating doseresponse relationship in meta-analysis: methodology, examples, and comparison. PeerJ Preprints. 2018;6:e27277v1.

[10] Greenland S, Longnecker MP. Methods for trend estimation from summarized dose-response data, with applications to meta-analysis. American Journal of Epidemiology. 1992;135:1301–9.

[11] Liu X, Wang W, Tan Z, Zhu X, Liu M, Wan R, et al. The relationship between vitamin D and risk of atrial fibrillation: a dose-response analysis of observational studies. Nutrition journal. 2019;18:73.

[12] Chang TS, Ding Y, Freund MK, Johnson R, Schwarz T, Yabu JM, et al. Prior diagnoses and medications as risk factors for COVID-19 in a Los Angeles Health System. medRxiv. 2020.

[13] Merzon E, Tworowski D, Gorohovski A, Vinker S, Golan Cohen A, Green I, et al. Low plasma 25(OH) vitamin D level is associated with increased risk of COVID-19 infection: an Israeli population-based study. FEBS J. 2020.

[14] Higgins JP, Green S. Cochrane Handbook for Systematic Reviews of Interventions Version 5.2.0 [updated March 2017]The Cochrane Collaboration, 2017. Available from http://handbook.cochrane.org.chanpter: 10.4.3.1 Recommendations on testing for funnel plot asymmetry2011.

[15] D’Avolio A, Avataneo V, Manca A, Cusato J, De Nicolo A, Lucchini R, et al. 25-Hydroxyvitamin D Concentrations Are Lower in Patients with Positive PCR for SARS-CoV-2. Nutrients. 2020;12.

[16] Daneshkhah A, Agrawal V, Eshein A, Subramanian H, Roy HK, Backman V. The Possible Role of Vitamin D in Suppressing Cytokine Storm and Associated Mortality in COVID-19 Patients. medRxiv. 2020:2020.04.08.20058578.

[17] Carpagnano GE, Di Lecce V, Quaranta VN, Zito A, Buonamico E, Capozza E, et al. Vitamin D deficiency as a predictor of poor prognosis in patients with acute respiratory failure due to COVID-19. J Endocrinol Invest. 2020.

[18] Dror Y, Giveon SM, Hoshen M, Feldhamer I, Balicer RD, Feldman BS. Vitamin D levels for preventing acute coronary syndrome and mortality: evidence of a nonlinear association. The Journal of Clinical Endocrinology & Metabolism. 2013;98:2160–7.

[19] Hong Z, Tian C, Zhang X. Dietary calcium intake, vitamin D levels, and breast cancer risk: a dose-response analysis of observational studies. Breast cancer research and treatment. 2012;136:309–12.

[20] Shi H, Chen H, Zhang Y, Li J, Fu K, Xue W, et al. 25-Hydroxyvitamin D level, vitamin D intake, and risk of stroke: A dose-response meta-analysis. Clin Nutr. 2020;39:2025–34.

[21] Sassi F, Tamone C, D’Amelio P. Vitamin D: Nutrient, Hormone, and Immunomodulator. Nutrients. 2018;10.

[22] Martineau AR, Jolliffe DA, Hooper RL, Greenberg L, Aloia JF, Bergman P, et al. Vitamin D supplementation to prevent acute respiratory tract infections: systematic review and meta-analysis of individual participant data. BMJ (Clinical research ed). 2017;356:i6583.

[23] Liu X, Long C, Xiong Q, Chen C, Ma J, Su Y, et al. Association of angiotensin converting enzyme inhibitors and angiotensin II receptor blockers with risk of COVID-19, inflammation level, severity, and death in patients with COVID-19: A rapid systematic review and meta-analysis. Clin Cardiol. 2020.

[24] Teymoori-Rad M, Marashi SM. Vitamin D and Covid-19: From potential therapeutic effects to unanswered questions. Rev Med Virol. 2020:e2159.

[25] Jolliffe DA, Greenberg L, Hooper RL, Mathyssen C, Rafiq R, de Jongh RT, et al. Vitamin D to prevent exacerbations of COPD: systematic review and meta-analysis of individual participant data from randomised controlled trials. Thorax. 2019;74:337–45.

[26] Milam AJ, Furr-Holden D, Edwards-Johnson J, Webb B, Patton JW, Ezekwemba NC, et al. Are Clinicians Contributing to Excess African American COVID-19 Deaths? Unbeknownst to Them, They May Be. Health Equity. 2020;4:139–41.

[27] Khunti K, Singh AK, Pareek M, Hanif W. Is ethnicity linked to incidence or outcomes of covid-19? BMJ (Clinical research ed). 2020;369:m1548.

